# A Blood-Based Transcriptomic Algorithm and Scoring System for Alzheimer’s Disease Detection

**DOI:** 10.1101/2025.10.21.25338438

**Authors:** Prisha Goyal, Neha Arya

## Abstract

Alzheimer’s disease is a neurodegenerative disorder that affects more than 50 million people worldwide. Current diagnostic methods include cerebrospinal fluid testing and CT/MRI scans, which are either invasive, expensive, or non-specific. This study aimed to develop alternative diagnostic approaches for early detection by creating a transparent algorithm using blood-based transcriptomic biomarkers associated with Alzheimer’s. Towards this, a microarray dataset from NCBI GEO was obtained, which contained the expression levels of 14,113 genes in 180 subjects: 90 Alzheimer’s cases and 90 non-Alzheimer’s controls. The Mann–Whitney U Test and the Holm–Bonferroni *p*-value correction were applied for feature selection, yielding 8 statistically significant genes. Further, symbolic regression using the Quantum Lattice technique led to the generation of a 3-gene mathematical function that could be utilized for Alzheimer’s prediction. The three genes identified in the regression model were FBRSL1, TRIB2, and LY6G6D, the first two being novel discoveries. Thereafter, the results were converted to a 100-point scoring system intended for clinical use, and the diagnostic scores were validated using the testing dataset. The model assigned scores lower than 20 to non-Alzheimer’s controls with 92% accuracy and scores greater than 80 to Alzheimer’s patients with 75% accuracy. The scoring system is a successful proof of concept that can serve as a starting point for more accurate risk systems in the future, and the identified biomarkers can be further validated in other cohorts of Alzheimer’s patients.

## 1 Introduction

Alzheimer’s disease (AD) is a progressive neurodegenerative disorder characterized by cognitive decline and behavioral impairment [1]. It affects more than 50 million people worldwide and is predicted to rise to over 150 million in 2050 with the increasing elderly population. Two-thirds of individuals affected by this disease reside in low- and middle-income countries (LMICs). The World Alzheimer Report identified the diagnosis and treatment gap in LMICs to be 90%, due to reasons such as limited awareness and knowledge, access to diagnostic tools, healthcare system capacity, cultural and social stigma, and economic considerations [2]. This underscores the value of accessible and rapid diagnostic methods for widespread impact [3]. Furthermore, earlier detection leads to more manageable disease trajectories and improved care outcomes [4].

Currently, clinicians employ a variety of diagnostic tests for the detection of Alzheimer’s in a patient. Neuropsychological tests, such as the Mini-Mental State Exam (MMSE) and the Montreal Cognitive Assessment (MoCA), assess a person’s cognitive abilities including memory, executive function, and reasoning [5]. These, and multiple other tests, are widely utilized due to their affordability and ease of administration for the clinical assessment of AD. However, while the MMSE has been found to adequately distinguish between individuals with and without dementia, its diagnostic accuracy diminishes in older adults with limited formal education, making it substantially less useful for patients in developing countries [6].

Cerebrospinal Fluid (CSF) testing is another diagnostic method used to detect biomarkers such as amyloid *β* (*Aβ*) and p-Tau. However, this procedure is highly invasive and is often impractical in resource-limited settings due its high cost [7].

Imaging tools such as CT (computed tomography) and MRI (magnetic resonance imaging) scans are commonly used in the detection of dementia. While both are available in under-resourced areas, measures such as hippocampal volume are not specific to Alzheimer’s and fail to give conclusive results [8, 9]. Fluorodeoxyglucose positron emission tomography (FDG-PET), though a reliable indicator of AD in assessing early-stage dementia, is not widely available and is relatively expensive [8].

Current machine learning models use blood-based biomarkers, which have recently gained significant attention as minimally invasive, scalable, and clinically feasible tools for Alzheimer’s disease detection, particularly in underdeveloped settings where advanced imaging or CSF collection is impractical [10]. The complexity of Alzheimer’s necessitates methods that can detect high-dimensional patterns in data and process vast gene expression datasets, establishing machine learning as a critical tool. Recent literature on Alzheimer’s disease diagnosis using transcriptomics data has employed a large variety of machine learning approaches. However, their accuracies typically range between 70% and 80% [11, 12, 13, 14], remaining low relative to other biological prediction tasks such as the detection of cancer or diabetes.

A study by Voyle et al. [11] combined the pathway-level gene expression analysis approach (PLAGE) with the Random Forest technique in order to develop a more robust diagnostic model. Using blood expression data from the AddNeuroMed (ANM) and Dementia Case Registry (DCR) studies, models based on only demographic data, demographic and gene level data, and demographic and PLAGE score data were built. Both the PLAGE and gene level models resulted in a similar performance accuracy of 65.7%.

Rye et al. [12] used Partial Least Squares Regression (PLSR) to develop a machine learning classification algorithm for early AD detection. A 96-gene expression array was identified and validated using two tests. The first compared the diagnosis of the algorithm against a clinical diagnosis made at autopsy, while the second compared the algorithm against cerebrospinal fluid biomarker status. An accuracy of 71.6 ± 10.3% was achieved in the first test, and an agreement of 80% was found in the second.

Lunnon et al. [13] used microarray-based blood gene expression profiling with a Random Forest classifier to develop an AD diagnostic model. They used data from the AddNeuroMed cohort to train and validate a 48-gene classifier, which achieved 75% accuracy in distinguishing AD from controls. Its accuracy in comparison to a structural MRI-based classifier in the same individuals was slightly lower, with an 85% accuracy for MRI. Combining both modalities led to an accuracy of 84%.

Abdullah et al. [14] found transcriptomics biomarkers for AD patients in Malaysia, using data from the Towards Useful Ageing (TUA) project. The dataset of 22,254 genes was narrowed to 68 genes using Boruta’s feature selection. Multiple classifiers were evaluated, with an elastic net logistic regression model performing best. It identified 16 potential biomarkers which had an accuracy of 81.59% and sensitivity of 85.19%.

Most of these studies, however, focus on binary classification models. This forces each sample into one of two classes, limiting the clinical interpretability of physician judgment based on granularity. Recent studies have emphasized the importance of interpretability in machine learning models for Alzheimer’s diagnosis. Alatrany et al. introduced explainable rule-based ML models using SHAP and LIME to identify key AD risk contributors like memory and judgment scores [15]. Emerging recommendations also favor simpler diagnostic models as shown by Diogo et al., who demonstrated that classifiers using minimal features such as hippocampal and frontal regions still achieve high predictive accuracy while improving generalizability across datasets [16].

The aim of this study was to identify differentially expressed genes from blood transcriptome data and determine the prediction accuracy of an algorithm and scoring system based on a minimal and feasible gene signature in detecting Alzheimer’s disease. This study’s novelty is a scoring system that translates gene expression patterns into a 0–100 risk scale, offering clinicians a probabilistic estimate that enables more flexible interpretation. Additionally, the model utilizes a transparent symbolic regression algorithm, making it feasible for deployment.

## 2 Materials & Methods

### 2.1 Dataset Source, Subjects, Data Collection, and Study Design

Through the NCBI Gene Expression Omnibus repository (GEO), a publicly available, anonymized dataset was retrieved (accession number GSE85426) which contained the preprocessed and normalized gene expression profiles of 180 individuals, 90 of which had Alzheimer’s disease while the other 90 were controls without Alzheimer’s or dementia [17].

The data was collected by utilizing microarray platforms to quantify relative abundance of messenger RNA (mRNA) within a sample. A microarray works by using complementary oligonucleotide probes that bind only to specific genes. Using microarrays, it is possible to find expression levels for thousands of genes simultaneously [18].

This study employed a retrospective in silico design using an existing NCBI GEO dataset and employed computational and statistical methods to analyze gene expression data to develop a diagnostic scoring system for Alzheimer’s disease.

### 2.2 Feature Selection and Computational Tools

The independent variables were the gene expression levels, a quantitative continuous variable. The dependent variable was whether the patient’s diagnosis was Alzheimer’s or non-Alzheimer’s, a categorical variable. The dataset was split using a 50-50 train-test ratio since symbolic regression is not hampered by small training datasets and a larger test set was desired [19]. In order to find the best-fit equation with the least number of gene signatures out of 14,113 genes, a systematic analytical pipeline was implemented, as depicted in Figure 1.

**Figure 1:**
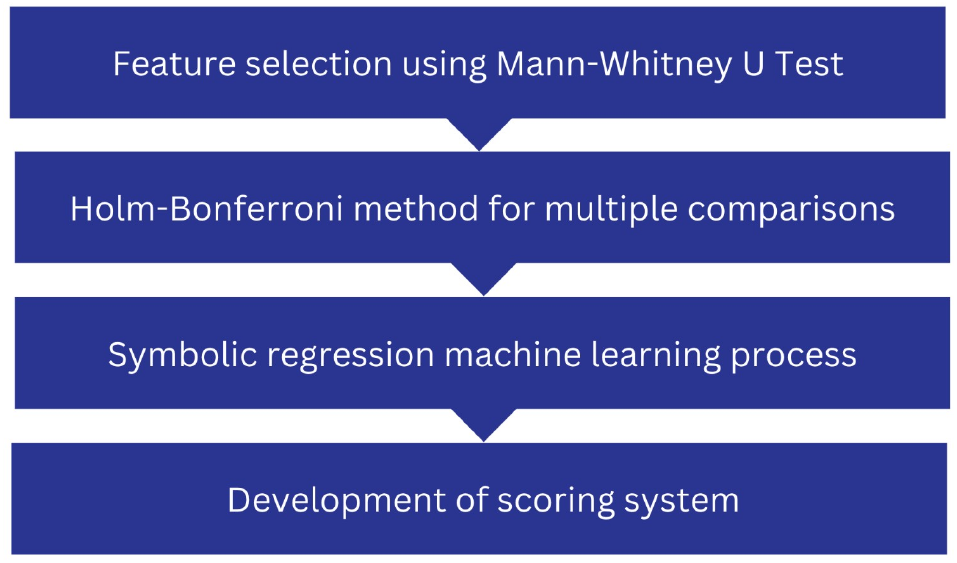
Flowchart outlining the overall research design. Feature selection on data was done using statistical tests, symbolic regression was used for model building, and a clinically interpretable scoring system for Alzheimer’s disease detection was developed

The Mann-Whitney U Test, which is the non-parametric alternative to the independent sample t-test, was performed on expression level distributions for each gene. This test evaluates statistical differences in gene expression distributions between AD and non-AD samples and is suitable for non-parametric data, such as gene expression levels [20]. Next, the Holm-Bonferroni Method for Multiple Comparisons was applied. The equation below shows Holm’s formula, which is used to calculate an adjusted alpha for the first-ranked p-value [21]. It adjusts significance thresholds in multiple hypothesis testing to reduce the likelihood of false positives when identifying differentially expressed genes between Alzheimer’s and non-Alzheimer’s subjects.

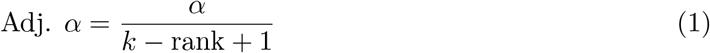

After applying these two tests, 8 statistically significant differentially expressed genes (DEGs) were discovered and used to create a reduced and more efficient search space that could be more thoroughly searched.

All computational analyses were conducted in Python, using numpy for numerical processing, pandas for data storage, seaborn for visualization of violin plots, sklearn for train-test splitting, scipy for applying the Mann-Whitney U Test, and Feyn for training the quantum lattice.

### 2.3 Model Building

Symbolic regression is a modeling approach that derives mathematical expressions to optimally describe input-output relationships for a given dataset. It was conducted using the Quantum Lattice, which derives mathematical models through simulated exploration of a finite multi-dimensional lattice space. Model selection was guided by the Bayesian Information Criterion (BIC) to balance fit and parsimony. The probability distribution initially starts as a uniform distribution, and is adjusted for the next generation to favor the model structure of the best models in the previous generation [22]. The symbolic regression model was trained over 20 epochs with the Bayesian Information Criterion as its loss function.

### 2.4 Statistical Tools

Parametric tests are conventionally used in hypothesis testing studies. However, they have strict assumptions that are often unmet by real-world health data: multivariate normality, the assumption that the data follows a normal distribution; random sampling, the assumption that the data comes from a random and representative sample; and homoscedasticity, the assumption that the variance is the same across independent variables. These conditions are often assumed to be true, resulting in oversimplified models that may not adequately capture the complexity of the data. Non-parametric tests, on the other hand, make fewer assumptions about the data, enhancing their reliability [21, 23].

In this study, the Mann-Whitney U Test was used for identifying statistically significant genes due to its non-parametric nature. The Holm-Bonferroni correction was then applied to control for Type I errors resulting from multiple hypothesis testing.

## 3 Results

The results of the analysis are summarized in the pipeline diagram shown in Figure 2A. Following feature selection, eight genes were found to be significantly differentially expressed between Alzheimer’s disease (AD) and non-AD subjects. These genes served as inputs to the symbolic regression model, which ultimately selected FBRSL1 (downregulated), TRIB2 (upregulated), and LY6G6D (downregulated) as the best predictors of AD status. All three genes demonstrated strong statistical significance with *p*-values < 0.01. The violin plots of the three genes are shown in Figure 2B.

**Figure 2.**
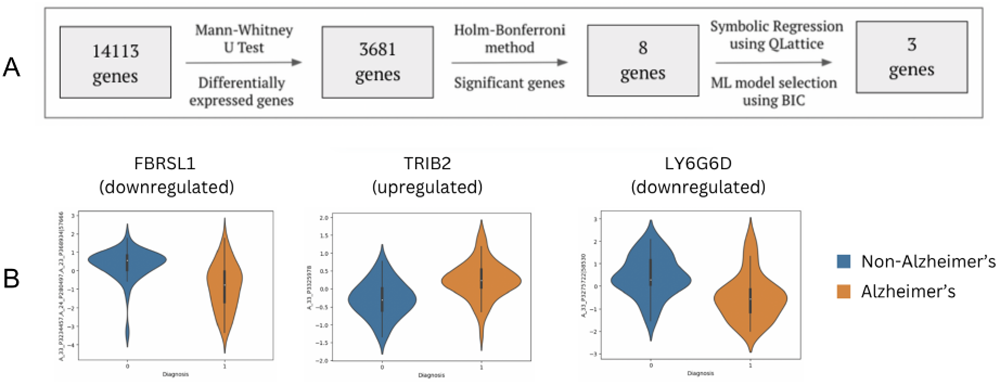
A: Stepwise results of the analysis pipeline, starting with 14113 genes, identifying 3681 differentially expressed genes, narrowing to 8 statistically significant genes, and ultimately identifying a 3-gene signature (FBRSL1, TRIB2, LY6G6D) through symbolic regression. B: Violin plots displaying the distribution of expression levels for the three key biomarker genes (FBRSL1, TRIB2, and LY6G6D) in Alzheimer’s patients compared to non-Alzheimer’s controls.

The symbolic regression Quantum Lattice model generated a mathematical equation based on these three genes (Figure 5A), which produced raw output values reflecting AD risk.

The QLattice symbolic regression model’s output predictions are adjusted by another equation to be more interpretable to clinicians, generating a score from 0 to 100. The equation that maps the model predictions to the scoring system is shown below, and a graphical representation of the correspondence between the prediction (on the x-axis) and scores (on the y-axis) is shown in Figure 3B. The model’s predictions are clustered towards the extremes. The scoring function distributes them evenly across 0 to 100, a more interpretable range for clinicians. Within the full range, the scores are classified into three buckets as detailed in Figure 3B.

**Figure 3.**
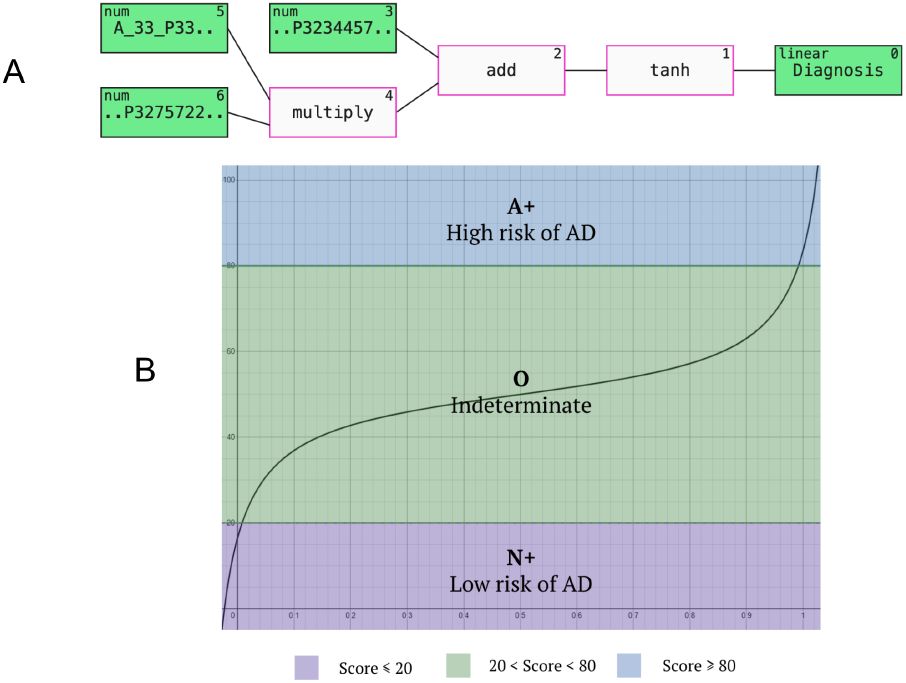
A: Mathematical relationship between the three biomarker genes (FBRSL1, TRIB2, LY6G6D) and Alzheimer’s disease risk, generated using the QLattice technique. B: Graphical mapping between the raw model predictions (x-axis) and the transformed clinician scoring system (y-axis), showing how the continuous outputs are distributed into a standardized 0–100 scale, with thresholds for high, medium, and low Alzheimer’s risk categories.

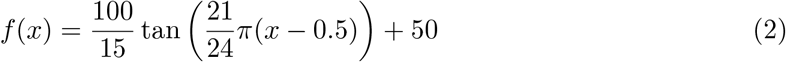

To assess the performance of the risk scoring system, the equation was applied to 90 test cases, 45 Alzheimer’s and 45 non-Alzheimer’s cases. Out of 12 cases scoring 20 or less, 11 (92%) were accurately identified as non-Alzheimer’s cases. Similarly, out of 20 cases having a score of 80 or more, 15 (75%) were accurately found to have Alzheimer’s disease. The results are summarized in Table 1.

**Table 1:** Validation of scoring system’s predictions on the test dataset. Scores *≤* 20 indicate low probability of Alzheimer’s and scores *≥* 80 indicate high probability.

| Range of Score | Subjects correctly predicted by scoring system | Probability of AD |
| --- | --- | --- |
| N+ (Score $\leq 20$) | 11/12 patients (92% accuracy) | Low |
| A+ (Score $\geq 80$) | 15/20 patients (75% accuracy) | High |

This study created an accurate scoring system that uses only a 3-gene signature, including two novel genes, to score patients between 0 and 100. A score of zero means a very low probability, and 100 means a high probability of Alzheimer’s disease. When the model’s predictions fall into the upper quartile (score *≥* 80) or lower quartile (score *≤* 20), it correctly differentiates between patients with or without Alzheimer’s while also providing physicians with a risk score for every suspected case to inform their own clinical judgment.

## 4 Discussion

This study aimed to develop a clinically interpretable diagnostic model for Alzheimer’s disease using blood-based transcriptomic data. By leveraging symbolic regression and a minimal 3-gene panel, the model produced a 0-100 scoring system which correctly predicted non-AD and AD cases with 92% accuracy in the lower quartile and 75% accuracy in the upper quartile.

These findings demonstrate improved performance relative to prior studies on blood-based Alzheimer’s diagnosis using binary classifiers, which are shown in Table 2. The study by Voyle et al. utilized the AddNeuroMed cohort and achieved an accuracy of 69% [11]. The 96-gene test for Alzheimer’s developed in Norway by Rye et al. had an accuracy of 72% [12], while Lunnon et al. obtained an accuracy of 75% using 48 genes [13]. Better results were obtained by Abdullah et al. [14] with an accuracy of 82% using 16 genes. This model demonstrates competitive diagnostic performance with only three genes, enhancing simplicity and cost-effectiveness for clinical implementation.

**Table 2:** Comparison of gene signature sizes and their corresponding accuracies in different studies.

| Study | Size of gene signature | Accuracy |
| --- | --- | --- |
| Voyle et al. (2016) [11] | n/a | 69% |
| Rye et al. (2011) [12] | 96 genes | 72% |
| Lunnon et al. (2013) [13] | 48 genes | 75% |
| Abdullah et al. (2022) [14] | 16 genes | 82% |

Beyond performance, the biological plausibility of our gene panel further strengthens its relevance. While LY6G6D has been previously identified by Abdullah et al. as an AD biomarker [14], the other two genes are novel discoveries with high plausibility as Alzheimer’s markers. FBRSL1 (Fibrosin-Like 1) has been associated with delayed memory recall in a study by Schuur [24]. While not previously validated as an AD marker, its connection to cognitive function makes it a strong candidate for further investigation. TRIB2 (Tribbles Pseudokinase 2) has been characterized in a recent review article by Guo et al. [25] as a protein of high interest in Alzheimer’s research. The review highlighted TRIB2 as a critical regulator of T cell naivety, with its decline contributing to age-related naïve/memory T cell imbalance. This study reinforces the likelihood of this previously suggested biomarker’s contribution to AD pathophysiology, which is good confirmation that its pattern-matching is aligned with established mechanisms. This further supports the investigation of FBRSL1 as a candidate biomarker, as it might turn out to be a comparably strong candidate to LY6G6D and TRIB2.

In contrast to other studies that employ larger panels, this study uses a limited 3-gene panel (FBRSL1, TRIB2, and LY6G6D), making it simple, affordable, and scalable for broad use. In contrast to conventional machine learning techniques such as random forests or SVMs, this study uses symbolic regression, generating a transparent and interpretable mathematical model that physicians can readily assess. The scoring system used here is not a conventional black-box machine learning algorithm, but a transparent mathematical equation that can be scrutinized and modified [26]. Symbolic regression has been used in physics to create formulas that model physical phenomena [27], and that practice was translated in this study to a neurological use case. Such transparent AI models drive translational research in medicine, and their innovative scoring system helps clinicians make diagnoses. Such transparency enhances interpretability and fosters clinical trust, which is necessary for regulatory approval and large-scale adoption by clinicians [28].

The limitations of this study include the relatively small size of the dataset, which could be remedied by obtaining a larger training dataset, yielding more generalized findings. The dataset also only contained patient data from a limited geographic region, which potentially limits the generalizability of the results. The scoring system was only validated internally, and the simplicity of the 3-gene signature may have affected the overall diagnostic accuracy.

In the future, obtaining larger sample sizes could enhance the diversity of the data, allowing for more generalizable models that overfit less and produce more accurate predictions. The results obtained in this study should also be further validated. The machine learning model and scoring system should be validated on an external dataset of Alzheimer’s and control patients. A user-friendly website or app could be developed that handles the prediction and conversion to point score for the clinician. Perhaps more importantly, TRIB2 and FBRSL1 warrant further investigation through in vivo studies, genome-wide association analyses, and other laboratory experiments to understand their mechanistic roles in the pathophysiology of Alzheimer’s disease and their potential as diagnostic or prognostic biomarkers or therapeutic targets.

## 5 Conclusion

A 3-gene signature using statistically significant transcriptomic blood biomarkers was identified in this study, including two novel biomarkers warranting further investigation and validation. The biomarker panel is suitable for implementation in public health contexts, especially in developing countries. The clinically useful scoring system that was developed is a proof-of-concept that could catalyze the development of more robust diagnostic systems by using larger datasets and combining gene panels with radiological findings and already identified biomarkers found in blood, such as amyloid-beta and tau proteins, to help in the early detection of Alzheimer’s disease. This approach could facilitate monitoring of Alzheimer’s disease progression in a patient. The transparent machine learning algorithm created using symbolic regression establishes a foundation for the development of clinically reliable models to gain regulatory approval. In summary, this study introduces a promising blood-based transcriptomic approach for AD detection.

## Data Availability

All data used in the study are openly available online at the NCBI Gene Expression Omnibus (GEO) database under accession number GSE85426 at https://www.ncbi.nlm.nih.gov/geo/query/acc.cgi?acc=GSE85426.

## 6 Acknowledgements

The authors would like to acknowledge the contributors of the dataset used in this study—Samsudin AZ, Abdul Majeed A, and Ramasamy K—along with their affiliated institution, University Teknologi Mara, for making this dataset, “Peripheral blood gene expression as a biomarker for early detection of Alzheimer’s disease” (accession number GSE85426), publicly accessible on the National Center for Biotechnology Information (NCBI) Gene Expression Omnibus (GEO) platform.

PG thanks the Initiative for Research and Innovation in STEM for their guidance and additional mentorship.

NA would like to acknowledge AIIMS Bhopal for support.

## References

[1] Alzheimer’s disease - Symptoms and causes. (2024, December 27). Retrieved from https://www.mayoclinic.org/diseases-conditions/alzheimers-disease/symptoms-causes/syc-20350447

[2] Prince, M., Bryce, R., Ferri, C., & Alzheimer’s Disease International. (2011). World Alzheimer Report 2011. In Alzheimer’s Disease International [Report]. https://www.alzint.org/u/WorldAlzheimerReport2011.pdf

[3] Hardy-Sosa, A., León-Arcia, K., Llibre-Guerra, J. J., Berlanga-Acosta, J., Baez, S. d. l. C., Guillen-Nieto, G., & Valdes-Sosa, P. A. (2022). Diagnostic Accuracy of Blood-Based Biomarker Panels: A Systematic Review. Front. Aging Neurosci., 14, 683689. doi: 10.3389/fnagi.2022.683689

[4] Kavitha C, Mani V, Srividhya SR, Khalaf OI and Tavera Romero CA (2022) Early-Stage Alzheimer’s Disease Prediction Using Machine Learning Models. Front. Public Health 10:853294. doi: 10.3389/fpubh.2022.853294

[5] Wang, G., Estrella, A., Hakim, O., Milazzo, P., Patel, S., Pintagro, C., … Li, W. (2022). Mini-Mental State Examination and Montreal Cognitive Assessment as Tools for Following Cognitive Changes in Alzheimer’s Disease Neuroimaging Initiative Participants. J. Alzheimers Dis., 90(1), 263–270. doi: 10.3233/JAD-220397

[6] Spencer, R. J., Wendell, C. R., Giggey, P. P., Katzel, L. I., Lefkowitz, D. M., Siegel, E. L., & Waldstein, S. R. (2013). Psychometric limitations of the mini-mental state examination among nondemented older adults: an evaluation of neurocognitive and magnetic resonance imaging correlates. Exp. Aging Res., 23875837. Retrieved from https://pubmed.ncbi.nlm.nih.gov/23875837

[7] D’Abramo, C., D’Adamio, L., & Giliberto, L. (2020). Significance of Blood and Cerebrospinal Fluid Biomarkers for Alzheimer’s Disease: Sensitivity, Specificity and Potential for Clinical Use.J. Pers. Med., 10(3), 116. doi: 10.3390/jpm10030116

[8] van Oostveen, W. M., & de Lange, E. C. M. (2021). Imaging Techniques in Alzheimer’s Disease: A Review of Applications in Early Diagnosis and Longitudinal Monitoring. Int. J. Mol. Sci., 22(4), 2110. doi: 10.3390/ijms22042110

[9] Frisoni, G. B. (2001). Structural imaging in the clinical diagnosis of Alzheimer’s disease: problems and tools. J. Neurol. Neurosurg. Psychiatry, 70(6), 711–718. doi: 10.1136/jnnp.70.6.711

[10] Jia, L., Zhu, M., Yang, J., Pang, Y., Wang, Q., Li, Y., … Wei, Y. (2021). Prediction of P-tau/Aβ42 in the cerebrospinal fluid with blood microRNAs in Alzheimer’s disease. BMC Med., 19(1), 1–15. doi: 10.1186/s12916-021-02142-x

[11] Voyle, N., Keohane, A., Newhouse, S., Lunnon, K., Johnston, C., Soininen, H., … Arendash, G. (2015). A Pathway Based Classification Method for Analyzing Gene Expression for Alzheimer’s Disease Diagnosis. J. Alzheimers Dis., 49(3), 659–669. doi: 10.3233/JAD-150440

[12] Rye, Phil. D., Booij, B. B., Grave, G., Lindahl, T., Kristiansen, L., Andersen, H.-M., … Lönneborg, A. (2011). A Novel Blood Test for the Early Detection of Alzheimer’s Disease. J. Alzheimers Dis., 23(1), 121–129. doi: 10.3233/JAD-2010-101521

[13] Lunnon, K., Sattlecker, M., Furney, S. J., Coppola, G., Simmons, A., Proitsi, P., … Hodges, A. (2013). A Blood Gene Expression Marker of Early Alzheimer’s Disease. J. Alzheimers Dis., 33(3), 737–753. doi: 10.3233/JAD-2012-121363

[14] Abdullah, M. N., Wah, Y. B., Abdul Majeed, A. B., Zakaria, Y., & Shaadan, N. (2022). Identification of blood-based transcriptomics biomarkers for Alzheimer’s disease using statistical and machine learning classifier. Inf. Med. Unlocked, 33, 101083. doi: 10.1016/j.imu.2022.101083

[15] Alatrany, A.S., Khan, W., Hussain, A. et al. An explainable machine learning approach for Alzheimer’s disease classification. Sci Rep 14, 2637 (2024). 10.1038/s41598-024-51985-w

[16] Diogo, V.S., Ferreira, H.A., Prata, D. et al. Early diagnosis of Alzheimer’s disease using machine learning: a multi-diagnostic, generalizable approach. Alz Res Therapy 14, 107 (2022). 10.1186/s13195-022-01047-y

[17] GEO Accession viewer. (2016, August 10). Retrieved from https://www.ncbi.nlm.nih.gov/geo/query/acc.cgi?acc=GSE85426

[18] Dufva, M. (2009). Introduction to Microarray Technology. DNA Microarrays for Biomedical Research. Humana Press. doi: 10.1007/978-1-59745-538-1_1

[19] Keren, L. S., Liberzon, A., & Lazebnik, T. (2023). A computational framework for physics-informed symbolic regression with straightforward integration of domain knowledge. Sci. Rep., 13(1249), 1–17. doi: 10.1038/s41598-023-28328-2

[20] de Torrenté, L., Zimmerman, S., Suzuki, M., Christopeit, M., Greally, J. M., & Mar, J. C. (2020). The shape of gene expression distributions matter: how incorporating distribution shape improves the interpretation of cancer transcriptomic data. BMC Bioinf., 21(21), 1–18. doi: 10.1186/s12859-020-03892-w

[21] Troyanskaya, O. G., Garber, M. E., Brown, P. O., Botstein, D., & Altman, R. B. (2002). Nonparametric methods for identifying differentially. Bioinformatics, 18(11), 1454–1461. doi: 10.1093/bioinformatics/18.11.1454

[22] Bharadi, V. (2021). QLattice Environment and Feyn QGraph Models—A New Perspective Toward Deep Learning. Emerging Technologies for Healthcare. John Wiley & Sons, Ltd. doi: 10.1002/9781119792345.ch3

[23] Giacalone, M., Agata, Z., Cozzucoli, P. C., & Alibrandi, A. (2018). Bonferroni-Holm and permutation tests to compare health data: methodological and applicative issues. BMC Med. Res. Method., 18(1), 1–9. doi: 10.1186/s12874-018-0540-8

[24] Schuur, M. (2010, May 12). Genetic Determinants of Cognitive Function and Age-Related Brain Changes. Retrieved from https://repub.eur.nl/pub/19462

[25] Guo, L., Li, X., Gould, T., Wang, Z.-Y., & Cao, W. (2023). T cell aging and Alzheimer’s disease. Front. Immunol., 14, 1154699. doi: 10.3389/fimmu.2023.1154699

[26] Broløs, K. R., Machado, M. V., Cave, C., Kasak, J., Stentoft-Hansen, V., Batanero, V. G.,… Wilstrup, C. (2021). An Approach to Symbolic Regression Using Feyn. arXiv, 2104.05417. Retrieved from https://arxiv.org/abs/2104.05417v1

[27] Udrescu, S.-M., & Tegmark, M. (2020). AI Feynman: A physics-inspired method for symbolic regression. Sci. Adv., 6(16). doi: 10.1126/sciadv.aay2631

[28] Price, Nicholson W, II. (2015). Black-Box Medicine. Harvard Journal of Law & Technology, 28(2). http://jolt.law.harvard.edu/articles/pdf/v28/28HarvJLTech419.pdf

